# A Retrospective Study of Comparison Between Neostigmine Versus Sugammadex as Reversal Agent in Patients Posted for Interventional Pulmonolgy Procedures

**DOI:** 10.64898/2026.09.08.26362508

**Authors:** Sanjeeta Umbarkar, Malvika Patil, Harshada Patane, Prerana Dixit

**Author notes:** Contact number-9323273435 (Sanjeeta Umbarkar). Contact number-8668512396 (Malvika Patil).

## Abstract

**Background and aims:** Patients for interventional pulmonary procedures (IPP) are inherently high risk due to various comorbidities as well as the procedure being variable in duration and adverse events. Incomplete muscle reversal can delay rapid recovery and lead to adverse events in the postoperative period. Sugammadex as a reversal agent, may offer distinct advantages when residual neuromuscular blockade is poorly tolerated. Limited data is available on use of suggamadex in high risk IPP regaurding extubation times and adverse events. Studies on pulmonary complications and anaphylaxis in this subset of pulmonary patients is controvertial. Moreover in the Indian setup optimizing the turnaround time to justify the cost of suggamadex for IPP is yet to be determined. In this retrospective study we will compare the advantages of using Sugammadex over the conventional reversal agent Neostigmine for time to extubation and time to shifting the patient to PACU in patients undergoing IPP. The hemodynamic parameters and adverse events will also be noted.

**Materials and methods:** After ethics approval from institute (EC/OA-48/2025) we conducted a retrospective cohort study of 70 adults (18-60yrs) for a duration of 6 months in our institute who underwent IPP under general anaesthesia (tracheal dilation, tracheal biopsy, EBUS TBNA,BAL etc). They were categorized into two groups: Group A-suggamadex vs. Group B-neostigmine. TOF monitoring was recorded and time to extubation and shift to PACU was noted along with hemodynamics and adverse effects. The statistical software SPSS version 16 was used for the analysis.

**Results:** A total of 70 patients were included. Patients in Group sugammadex showed less time to extubate (5.80±0.96 vs 13.26±1.88, P value 0.0001) and discharge from PACU (10.49±1.44 vs 28.57±3.34 min, *P* = 0.000003) than in Group neostigmine. The difference between heart rate, blood pressure and respiratory rate and adverse events were not statistically significant.

**Conclusion:** Sugammadex shortens the time to extubate and discharge from PACU in patients undergoing IPP, accelerating recovery and turnaround time. No additional adverse events or hemodynamic instability were noted with suggamadex in this subset of patients. No coughing irritable airway.

## INTRODUCTION

Patients for interventional pulmonary procedures (IPP) are inherently high risk due to various comorbidities such as cancer, tuberculosis and other long standing lung disorders.^[1]^

Interventional pulmonary procedures such as tracheal dilatation, bronchial alveolar lavage, EBUS TBNA, foreign body aspiration, tracheal biopsy etc. are done with either fibreoptic bronchoscopy or rigid bronchoscopy.^[1]^ Bronchial bleeding, airway manipulation and varying length of these procedures make anaesthesia management challenging.^[1][2][3]^ Procedures can be done under MAC (monitored anaesthesia care), sedation using TIVA or under general anaesthesia. Ventilation strategies include jet ventilation, LMA (laryngeal mask airway) or endotracheal intubation.^[3]^

The anaesthesia goals for these patients is having minimum post-operative pulmonary dysfunction. [^2]^ This is achieved by using short acting muscle relaxants and a quick and rapid reversal and extubation.^[3][4]^ Neostigmine along with glycopyrolate is a standard reversal protocol in most institutes.^[3]^

Muscarinic side effects like increased secretions, bradycardia, nausea and vomiting and residual muscular blockade are some side effects of neostigmine which preclude its use in IPPs.

Suggamadex is a novel muscle reversal agent which doesn’t have the muscarinic side effects of neostigmine but has potential to cause allergic side effects and risk of anaphylaxis.^[4][5][6]^

But enough evidence does not exist for its use for IPPs and its benefit for post-operative pulmonary complications.^[4][5][6]^ Limited data is available on use of suggamadex in high risk IPP regarding extubation times and adverse events.^[2][3][4]^ Studies on pulmonary complications and anaphylaxis in this subset of pulmonary patients is controversial. Moreover in the Indian setup optimizing the turn around time to justify the cost of suggamadex for IPP is yet to be determined.^[4][5]^ In this retrospective study we will compare the advantages of using Sugammadex over the conventional reversal agent Neostigmine for time to extubation and time to shifting the patient to PACU in patients undergoing IPP. The hemodynamic parameters and adverse events will also be noted.

## AIM OF THE STUDY

In this retrospective study we will compare the advantages of using the novel reversal agent Sugammadex over the conventional reversal agent Neostigmine in terms of time to extubation and time to shifting the patient to recovery room in patients undergoing interventional pulmonology procedures.

## OBJECTIVES

- PRIMARY OBJECTIVE: To observe the time taken to extubation and time to shift of the Patient to PACU.
- SECONDARY OBJECTIVES:
  1. Note the hemodynamic alterations after administration of the reversal agent
  2. Note any adverse reaction to the reversal agent

## INCLUSION AND EXCLUSION CRITERIA

INCLUSION CRITERIA:

1. Age: 18-60 years
2. Patients posted for Interventional Pulmonology procedures under General Anaesthesia

We excluded patients who did not receive neuromuscular monitoring.

## MATERIALS AND METHODOLOGY

1. Our study duration was 6 months
2. Data collection: We compiled the data from the past medical records.
3. We then divided the data into 2 groups

Group A – those who were administered reversal agent Suggamadex

Group B – those who were administered reversal agent neostigmine.

From the data the following preoperative details of the patients were noted:

1. Patients demographic details
2. Preoperative assessement of patient – symptoms, comorbidities, examination findings, Other medications
3. Preoperative investigations of the patients

From the data the following intraoperative details of the patients were noted:

1. Baseline Heart rate, blood pressure, respiratory rate, saturation.
2. Any neuromuscular monitoring technique used.
3. Any technique for measuring depth of anesthesia.
4. Name of the procedure.
5. Duration of the procedure.
6. Type of anesthesia.
7. Presence or absence of shared airway between the Pulmonologist and the anesthesiologist.
8. The anaesthetic agents used for premedication, sedation, induction agent, maintenance agent and the neuromuscular blocking agent used.
9. The reversal agent given at the end of the procedure

From the data the following post-operative details of the patients were noted

1. TOF count, HR, BP, SPO2, RR before giving reversal, after giving reversal and at at 5 min, 10 min, 15 min, 30 min and 1 hour interval
2. TOF count before extubation
3. Time from administration of reversal agent till extubation
4. Time from administration of reversal agent till shift of the patient to the recovery
5. Any post-operative complications
6. And adverse reaction to the reversal agents

Here the criteria for time to extubation and time of shift of patient to recovery were Considered as follows:

- CRITERIA FOR EXTUBATION:
  1. TOF count 4
  2. TOF ratio > 0.9
  3. Return of spontaneous activity in form of eye opening, following commands, head lift, return of motor power and tone
  4. Stable hemodynamics
- CRITERIA FOR SHIFT OF PATIENT TO RECOVERY:
  1. Stable hemodynamics after extubation
  2. Presence of spontaneous activity in form of obeying commands, vocalizing, normal tone, power
  3. Absence of any adverse effects

PLACE OF STUDY: Department of Cardiovascular and Thoracic Anaesthesiology, Seth GS medical college and KEM hospital, Mumbai.

## SAMPLE SIZE

For calculating sample size, following was applied

Sample size was calculated using OPEEPI2.3.1 software, According to study by Lee T Y et al *, it was found that ppoFEV1 in Neostigmine group and Sugammadex group was 77.4±14.8 and 70.2±13.4 respectively. At 80% power and using Formula

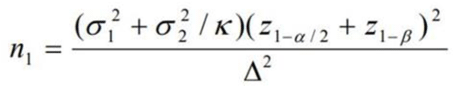

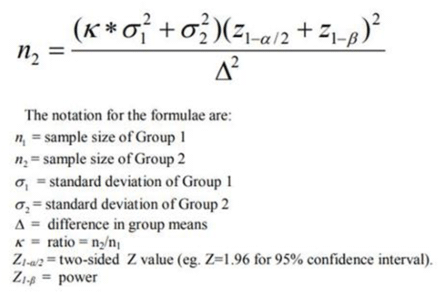

and comparing the means the sample size was calculated to be 35 in each group, with total sample size of 70.

## STATISTICAL ANALYSIS PLAN

The Data was collected, coded and entered in Microsoft excel 2016 and in order to display analytically, the results of this study, the following statistical

Method was adopted: – Continuous variables like time to extubation and shift to PACU, Age, Height, Weight, Heart Rate, Mean blood pressure and SPO2 etc. were expressed as Mean ± Standard Deviation and compared across the 2 groups using unpaired t test.

Categorical variables like number of with Adverse Events were expressed as number of patients and percentage of patients and compared across the 2 groups using Pearson’s Chi

Square test for Independence of Attributes. The statistical software SPSS version 16 was used for the analysis. An alpha level of 5% was considered, i.e. if any p value is less than 0.05 it will be considered as statistically significant.

## OBSERVATIONS-

From the collected data we observed that:

1. Thorough preoperative evaluation of the patients was done and investigations were checked.
2. Patients were divided in two groups: one who received Neostigmine and one who received Sugammadex
3. Baseline vitals (HR, BP, RR, SPO2, Baseline TOF count) were noted according to the standard ASA guidelines.
4. Patients were induced under General Anaesthesia using Inj Midazolam (0.03 mg/kg) iv Inj Fentanyl (2mcg/kg) iv Inj Propofol (2mg/kg) iv.
5. TOF(train of four) count was noted immediately before giving muscle relaxant. TOF count was 4.
6. Muscle relaxant Inj Rocuronium (1mg/kg)was given
7. Anaesthesia was maintained on Inhalational agent Sevoflurane titrated to acheieve MAC(minimum alveolar concentration) of 0.7 and top ups of Inj Rocuronium(0.1mg/kg)
8. TOF count 0 taken as deep plane of anaesthesia, muscle relaxant top up was given when TOF count reached 2.
9. At the end of the procedure TOF count was repeated. Once TOF count became 3 Reversal agent selected by the investigator was given in the following doses: Inj Sugammadex (2mg/kg) if group A and Inj Neostigmine (0.05mg/kg) if group B TOF count was noted immediately after giving relaxant, at 5 min, 10 min, 15 min, 30 min and 1 hour
10. Patient was extubated once the TOF count returned to 4 and TOF ration > 0.9
11. Time to extubation was noted (from the time of administration of reversal agent till time of extubation)
12. Time to shift the patient to recovery room was noted (from administration of reversal agent till shift of patient to recovery room
13. Hemodynamic trend was monitored to detect any adverse reaction to reversal agent On doing the statistical analysis of the of the data we observed the following:

In the current study, there is no significant difference between Sugammadex and Neostigmine with respect to changes in the heart rate. Both the drugs have near similar changes in the heartrate as shown in the table 1.

**Figure 1:**
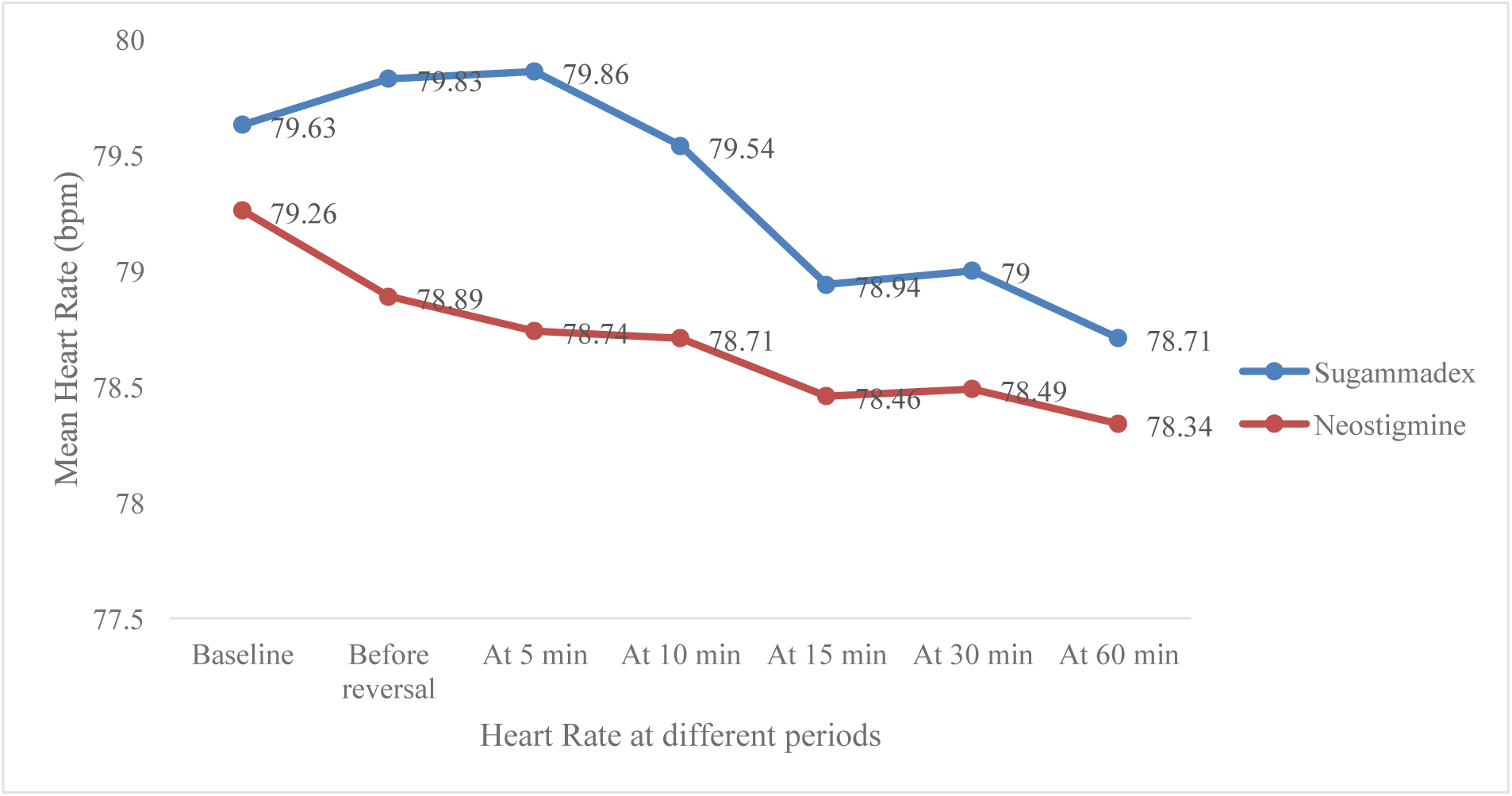
Graph showing the comparison between two drugs with changes in heart rate.

**Table 1:** Table showing the change in Heart Rate between the two drugs.

| Heart Rate (in bpm) | Sugammadex<br>N = 35<br>Mean±SD | Neostigmine<br>N = 35<br>Mean±SD | p value |
| --- | --- | --- | --- |
| Baseline | 79.63±11.26 | 79.26±12.71 | 0.8978 |
| Before reversal | 79.83±11.30 | 78.89±12.07 | 0.7376 |
| At 5 min | 79.86±10.95 | 78.74±11.67 | 0.6801 |
| At 10 min | 79.54±10.82 | 78.71±11.71 | 0.7950 |
| At 15 min | 78.94±10.89 | 78.46±11.19 | 0.8562 |
| At 30 min | 79.00±10.52 | 78.49±11.34 | 0.8459 |
| At 60 min | 78.71±10.25 | 78.34±11.28 | 0.8862 |

In the current study, there is no significant difference between Sugammadex and Neostigmine with respect to changes in the blood pressure. Both the drugs have near similar changes in the heartrate as shown in the table 2.

**Table 2:**
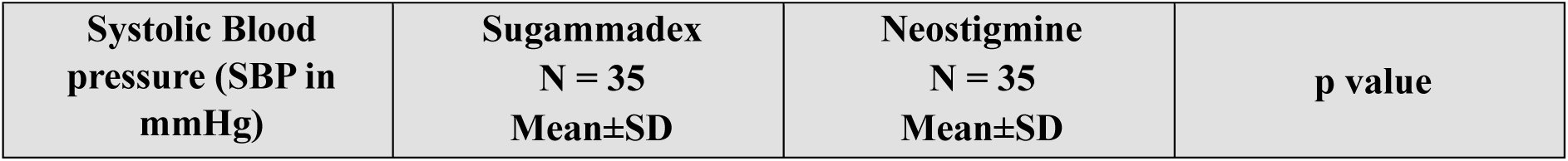
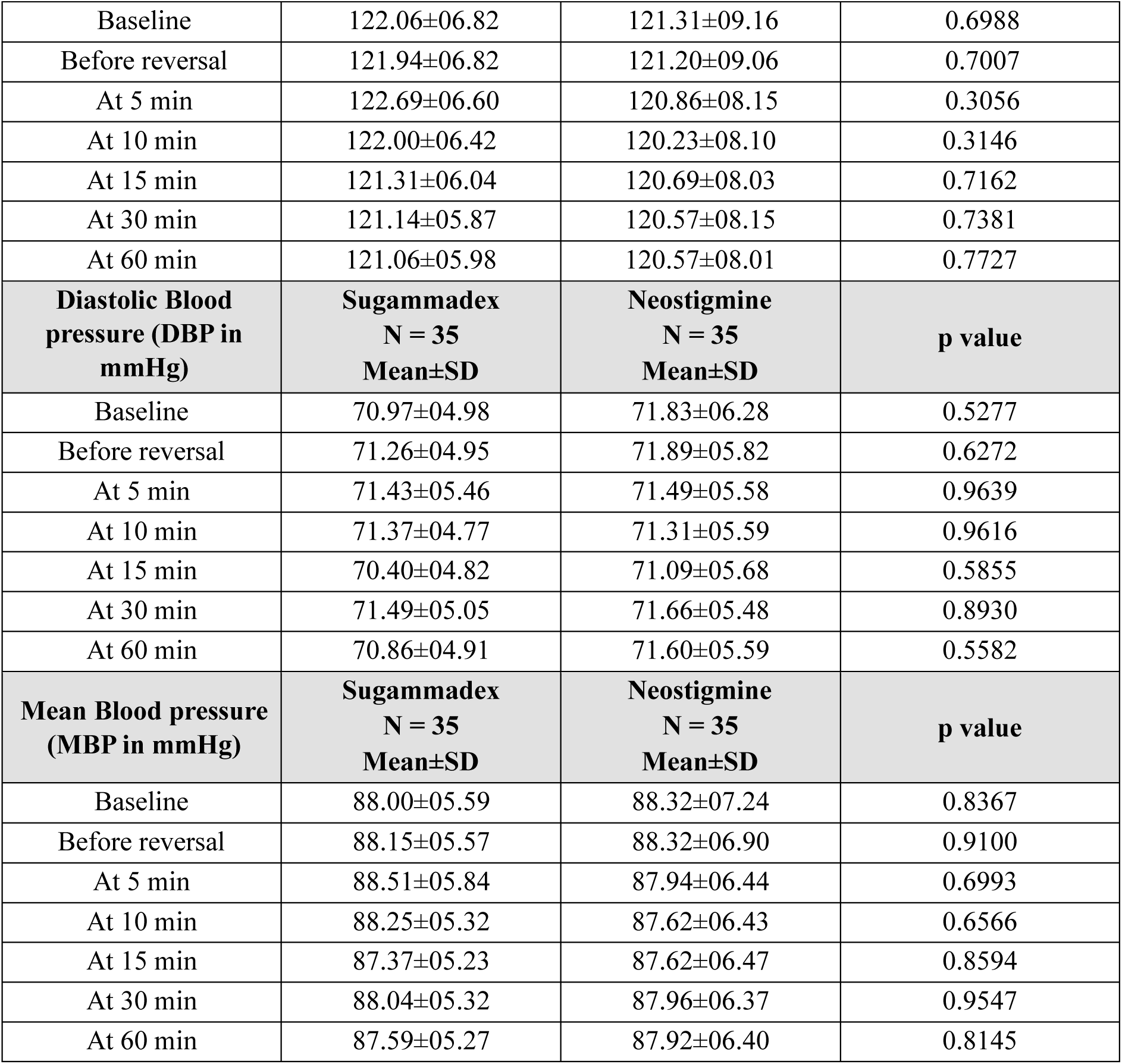
Table showing the change in blood pressure between the two drugs.

| Systolic Blood pressure (SBP in mmHg) | Sugammadex<br>N = 35<br>Mean±SD | Neostigmine<br>N = 35<br>Mean±SD | p value |
| --- | --- | --- | --- |
| Baseline | 122.06±06.82 | 121.31±09.16 | 0.6988 |
| Before reversal | 121.94±06.82 | 121.20±09.06 | 0.7007 |
| At 5 min | 122.69±06.60 | 120.86±08.15 | 0.3056 |
| At 10 min | 122.00±06.42 | 120.23±08.10 | 0.3146 |
| At 15 min | 121.31±06.04 | 120.69±08.03 | 0.7162 |
| At 30 min | 121.14±05.87 | 120.57±08.15 | 0.7381 |
| At 60 min | 121.06±05.98 | 120.57±08.01 | 0.7727 |
| **Diastolic Blood pressure (DBP in mmHg)** | **Sugammadex**<br>**N = 35**<br>**Mean±SD** | **Neostigmine**<br>**N = 35**<br>**Mean±SD** | **p value** |
| Baseline | 70.97±04.98 | 71.83±06.28 | 0.5277 |
| Before reversal | 71.26±04.95 | 71.89±05.82 | 0.6272 |
| At 5 min | 71.43±05.46 | 71.49±05.58 | 0.9639 |
| At 10 min | 71.37±04.77 | 71.31±05.59 | 0.9616 |
| At 15 min | 70.40±04.82 | 71.09±05.68 | 0.5855 |
| At 30 min | 71.49±05.05 | 71.66±05.48 | 0.8930 |
| At 60 min | 70.86±04.91 | 71.60±05.59 | 0.5582 |
| **Mean Blood pressure (MBP in mmHg)** | **Sugammadex**<br>**N = 35**<br>**Mean±SD** | **Neostigmine**<br>**N = 35**<br>**Mean±SD** | **p value** |
| Baseline | 88.00±05.59 | 88.32±07.24 | 0.8367 |
| Before reversal | 88.15±05.57 | 88.32±06.90 | 0.9100 |
| At 5 min | 88.51±05.84 | 87.94±06.44 | 0.6993 |
| At 10 min | 88.25±05.32 | 87.62±06.43 | 0.6566 |
| At 15 min | 87.37±05.23 | 87.62±06.47 | 0.8594 |
| At 30 min | 88.04±05.32 | 87.96±06.37 | 0.9547 |
| At 60 min | 87.59±05.27 | 87.92±06.40 | 0.8145 |

**Table 3:**
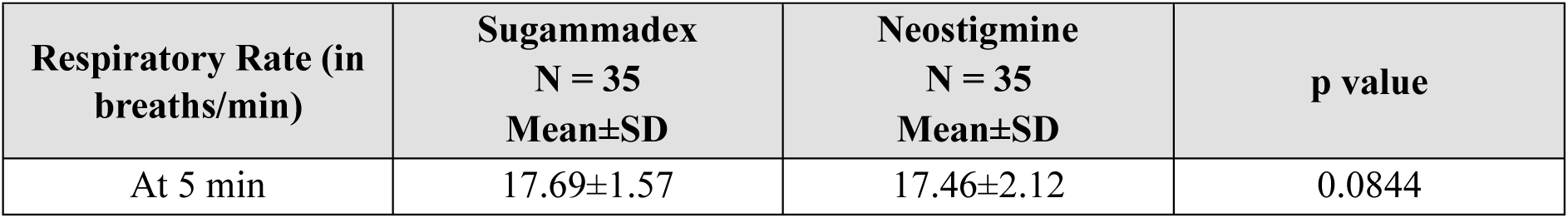
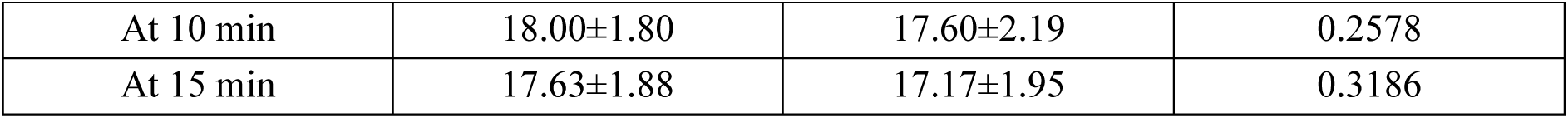
Table showing the change in Respiratory Rate between the two drugs.

There was no statistical difference between respiratory rate at 5 min, 10 min, 15 min after reversal in both the groups.

**Figure 2:**
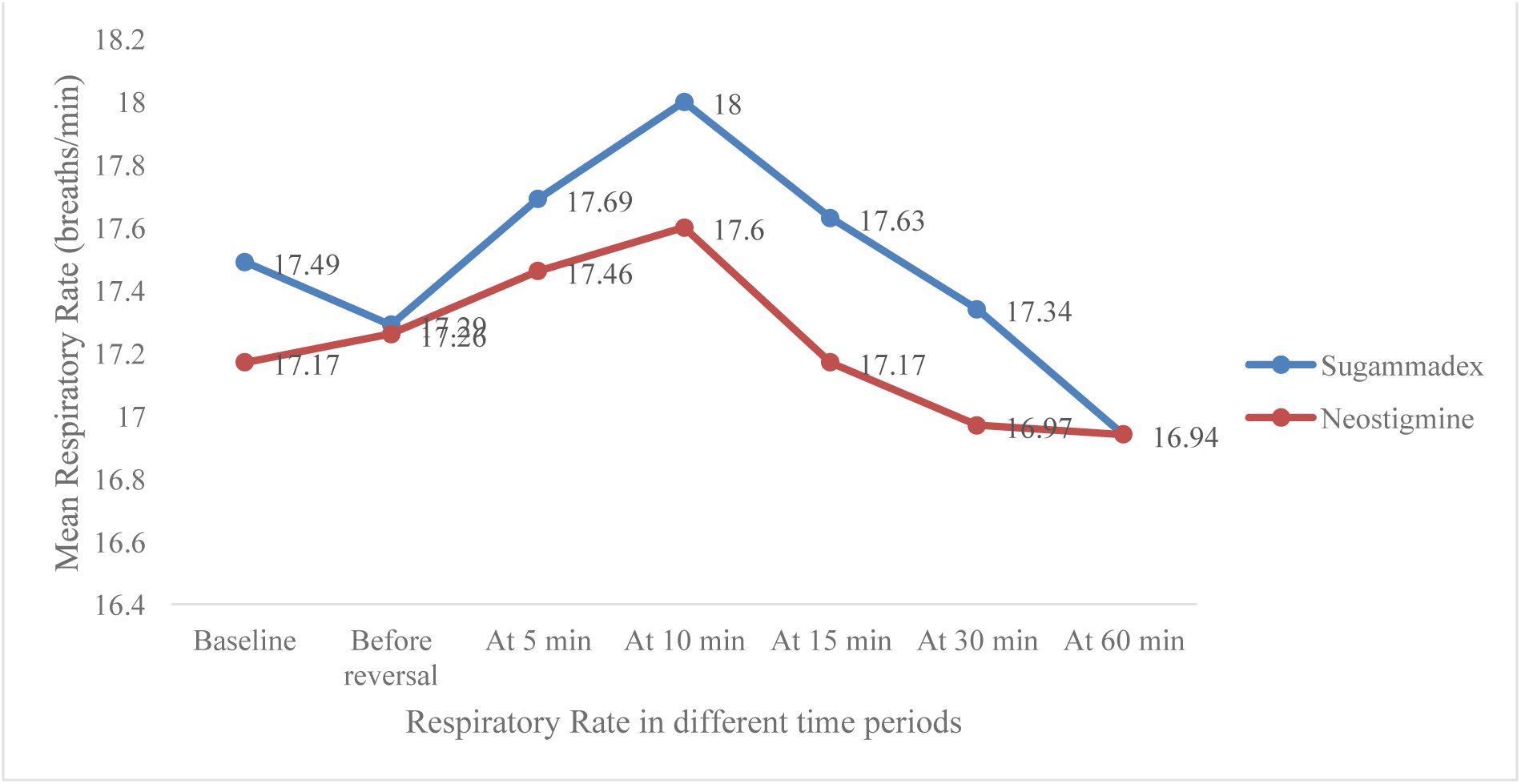
Graph showing the comparison of changes in respiratory rate between two drugs.

**Table 4:** Table showing the change in time to extubate and shift to recovery between the two drugs.

| Variables | Sugammadex<br>N = 35<br>Mean±SD | Neostigmine<br>N = 35<br>Mean±SD | p value |
| --- | --- | --- | --- |
| Time to extubate (in minutes) | 5.80±0.96 | 13.26±1.88 | **0.0001** |
| Time to shift to recovery (in minutes) | 10.49±1.44 | 28.57±3.34 | **0.000003** |

The current study showed that, patients who had been injected with Suggamadex had shorter period of extubation than neostigmine which had stronger association and was statistically significant. Similarly, shorter time was taken to shift the patients to recovery who had been injected with Suggamadex than neostigmine. This showed that, Suggamadex had better extubation and earlier shift to recovery after the effect of these drugs faded.

**Figure 3:**
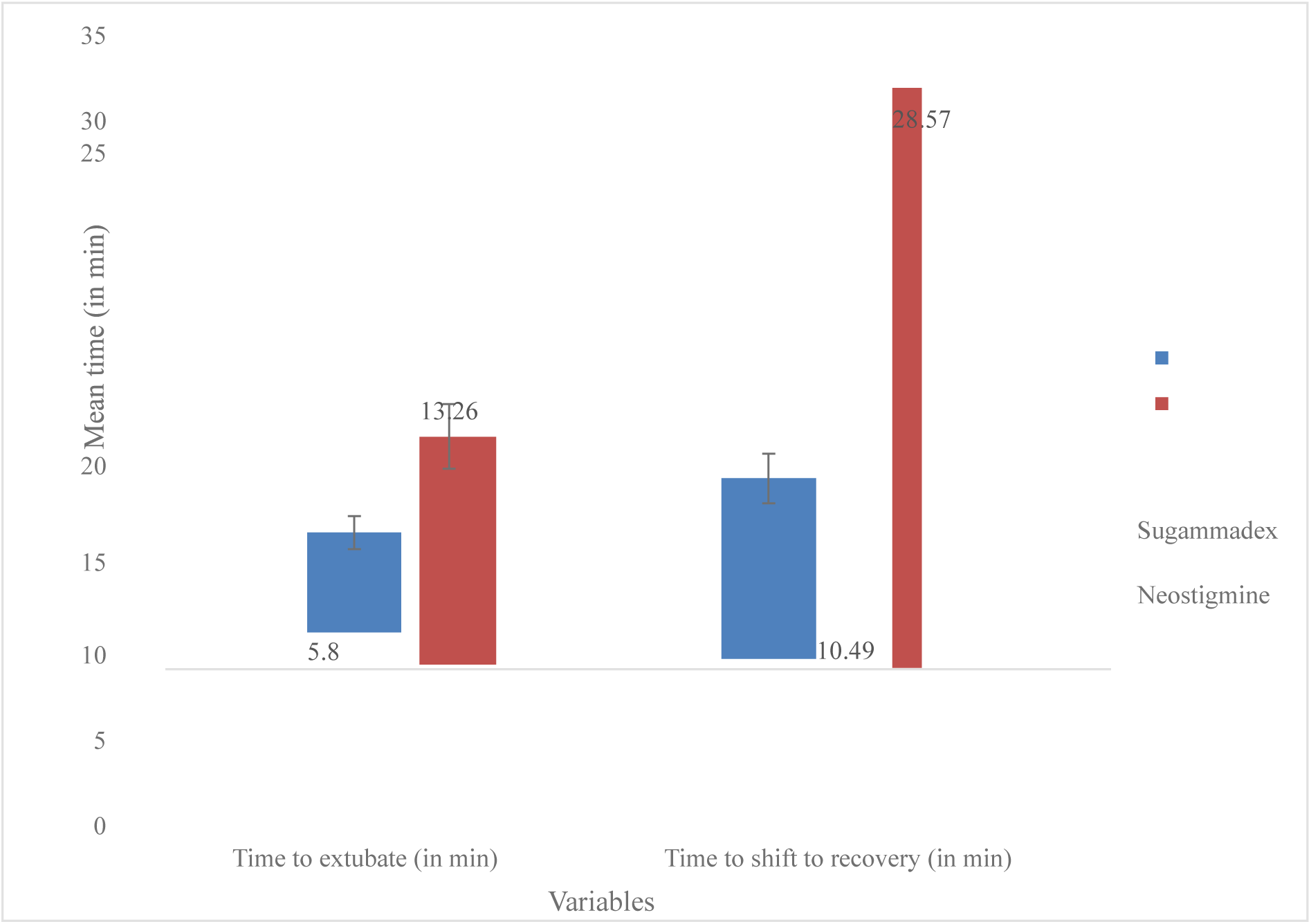
Graph showing the comparison between two drugs on extubation time and shifting to recovery.

**Table 5:** Table showing the TOF variations between two drugs.

| TOF | Sugammadex<br>N = 35<br>Mean±SD | Neostigmine<br>N = 35<br>Mean±SD | p value |
| --- | --- | --- | --- |
| Baseline | 4.00±0.00 | 4.00±0.00 | - |
| Before reversal | 3.00±0.00 | 3.00±0.00 | - |
| At 5 min | 4.00±0.00 | 3.49±0.51 | < 0.001 |
| At 10 min | 4.00±0.00 | 4.00±0.00 | - |
| At 15 min | 4.00±0.00 | 4.00±0.00 | - |
| At 30 min | 4.00±0.00 | 4.00±0.00 | - |
| At 60 min | 4.00±0.00 | 4.00±0.00 | - |

In the current study, patients with Sugammadex and Neostigmine had relatively similar TOF wherein, almost all patients had presence of 4^th^ twitch showing 0-5% paralysis. However, there was relatively better TOF in patients with Sugammadex than neostigmine in the initial 5 minutes period and later in 1 hour there were changes noted between two drugs with respect to TOF.

**Figure 4:**
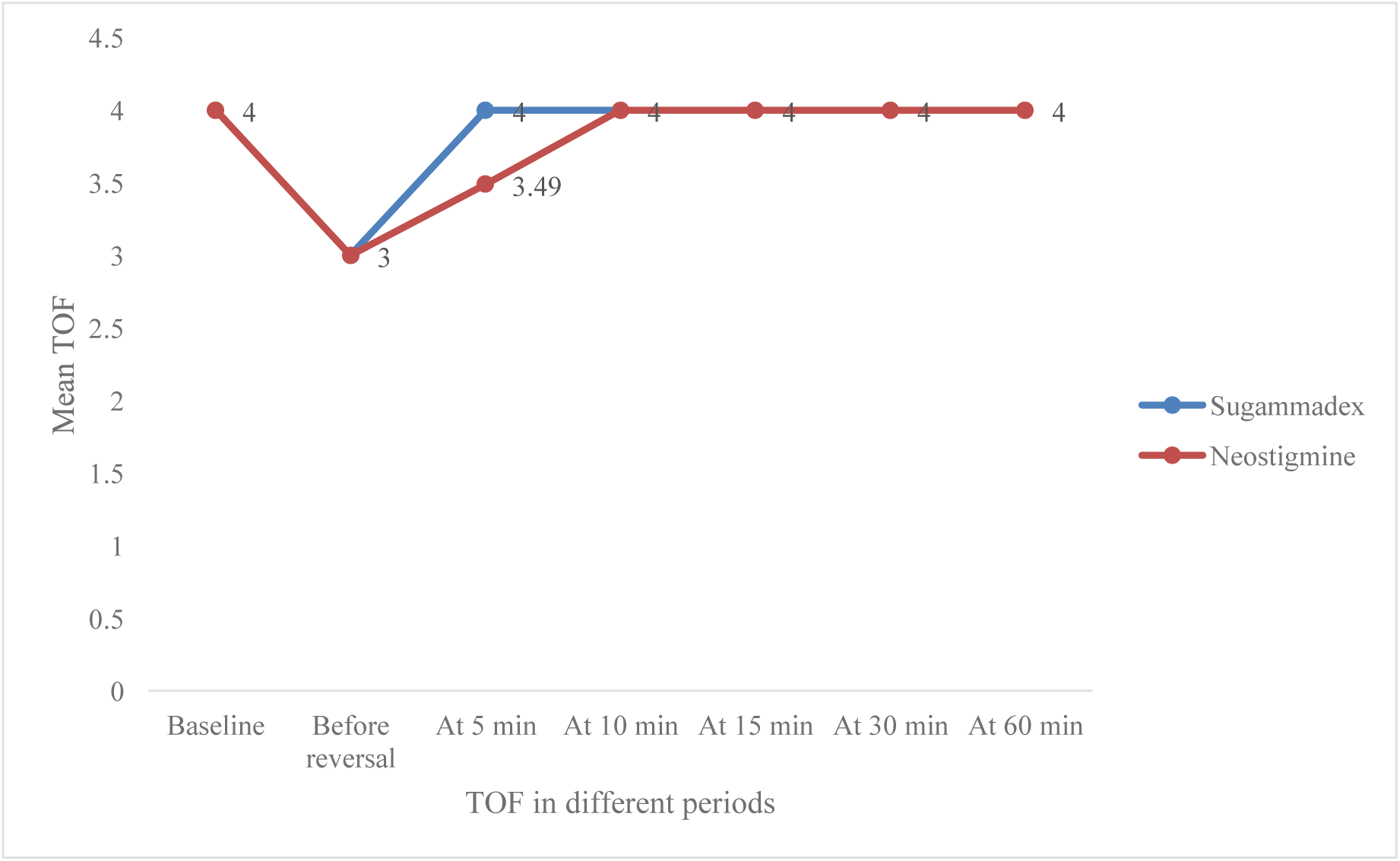
Graph showing the comparison between the two drugs with TOF.

**Table 6:** Table showing the comparison of adverse effects between two drugs.

| Adverse effects | Sugammadex<br>N = 35 | Neostigmine N<br>= 35 |
| --- | --- | --- |
| Yes | 00.00 (00.00) | 00.00 (00.00) |
| No | 35 (100.00) | 35 (100.00) |

There were no adverse effects noted in the current study in patients when compared both Sugammadex and neostigmine.

## RESULT

1. Heart rate variable: The p value across all the time periods is > 0.05. Thus, there is no statistically significant difference between the two drugs with respect to changes in the heart rate.
2. Blood pressure variable: The p value across all the time periods is > 0.05. thus, there is no statistically significant difference between the two drugs with respect to changes in the blood pressure.
3. Respiratory rate: In our study we found no relative changes in the respiratory rate at 5, 10 and 15 minutes post extubation (p value > 0.05)
4. Both time to extubation and time of shift of patient to recovery variables show p value < 0.05 I.e. 0.0001 and 0.000003 respectively showing statistically significant difference between the two groups.

From our study we found the following timings:

1. **Group Neostigmine**:
  A. **Time to extubation – 10 to 15 min**
  B. **Time to shift of patient to the recovery room – 20 to 30 min**
2. **Group Sugammadex**:
  **Time to extubation –4 min – 8 min**
  **Time to shift of patient to recovery – 8 – 10 min**

## SUMMARY

According to a study published in BMC Anesthesiology Sugammadex has been reported to lower the incidence of post operative residual neuromuscular blockade and other PPCs like pneumonia, atelectasis, NIV, reintubation as compared to Neostigmine but further evidence is required^(13)^ We conducted this study in accordance and to further support this evidence.

## DISCUSSION

Neostigmine and Sugammadex are both medications used to reverse the effects of neuromuscular blocking agents.^[1][2]3[4]^ Neostigmine works by inhibiting the enzyme acetylcholinesterase, leading to an increase in acetylcholine levels and ultimately reversing muscle paralysis. Sugammadex, on the other hand, works by forming a complex with the neuromuscular blocking agent, effectively removing it from the neuromuscular junction and rapidly reversing its effects.^[6][7][8]^ While Neostigmine is more commonly used due to its lower cost, Sugammadex is preferred in certain situations where a rapid and complete reversal of muscle paralysis is needed.

Neostigmine has a slower onset of action compared to Sugammadex, typically taking 5-10 minutes to reverse the effects of neuromuscular blockade. However, Neostigmine has a longer duration of action, lasting up to 60 minutes, which may be beneficial in cases where prolonged reversal is required. In contrast, Sugammadex has a rapid onset of action, with reversal occurring within 2-3 minutes, but its duration of action is shorter, lasting only 15-30 minutes.^[5][6][7][13][15][16]^

Neostigmine is associated with cholinergic side effects, such as bradycardia, bronchoconstriction, and gastrointestinal disturbances, due to its mechanism of action on the cholinergic system. In contrast, Sugammadex has a more favorable side effect profile, with minimal risk of cholinergic side effects.

However, Sugammadex has been associated with hypersensitivity reactions, such as anaphylaxis, in rare cases.^[3][4][5][13][14][16][17][18]^

Neostigmine is a more cost-effective option compared to Sugammadex, making it a preferred choice in settings where cost is a significant factor. However, the cost of Sugammadex may be justified in cases where rapid reversal of neuromuscular blockade is required, such as in emergency situations or when dealing with high-risk patients.^[9][10][13][^

From our study we can conclude that Sugammadex has a better profile in terms of quick, predictable extubation period which makes it a desirable drug for Interventional Pulmonology procedures as these patients have pre-existing pulmonary pathology, lower hypoxia threshold^(5)^shared airway between the pulmonologist and anesthesiologist^(4)^ and increased risk of bronchospasm. Risk of bronchospasm and anaphylaxis was similar in both groups.

There is no considerable difference in hemodynamic variability between the two agents.

Most of the pulmonary procedures are scheduled as day care procedures or where rapid turnaround time is required to fit multiple procedures in scheduled day. Suggamadex did considerably improve time to extubation and shift from PACU without compromising on patient safety.

We have observed that our patients had no post-operative pulmonary as well as other complications in the Sugammadex group. Sugammadex can be used in Enhanced recovery after surgery (ERAS) programs^(21)^

Many of our patients had pre-existing hyper reactive airways but no post-operative airway irritation or bronchospasm after suggamadex was seen.^[13]^

Suggamadex (dose) is also a favorable drug in cases of cannot intubate or cannot oxygenate (CICO) scenarios as patient can be quickly reversed when muscle relaxant rocuronium is used for induction. Neostigmine is not useful in these scenarios as no quick reversal is possible.^[9]^

Since Sugammaadex doesn’t cause muscarinic side effects it has a stable cardiovascular which makes it an ideal drug which can be used in patients having cardiac disease posted for cardiac as well as for non cardiac surgeries over neostigmine^(16)^.

The comfort of the patient, pulmonologist and the anaesthesiologist was better in the Sugammadex group.

Thus the cost of using suggamadex as an alternative to neostigmine is justified in this complex patient population of IPP for a better turn around time and safety profile.

## CONCLUSION

From our study we conclude that Sugammadex has better profile as compared to Neostigmine in terms of less time to extubation and shift of the patient to the recovery room in patients posted for interventional pulmonology procedures.

## LIMITATION OF THE STUDY

Since our study is of 6 months as part of curriculum in our 1 year course duration; to have clear idea about utility of Sugammadex in such procedures further prospective and metacentric studies are required.

## Data Availability

All data produced in the present study are available upon reasonable request to the authors

## ABBREVIATIONS

TOF: Train of four
ASA: American society of Anaesthesiology
PPCs: Post operative pulmonary complications
OT: Operation theatre
MAC: Minimum alveolar concentration
IPP: interventional pulmonary procedures.

